# Bayesian Variable Selection Utilizing Posterior Probability Credible Intervals

**DOI:** 10.1101/2021.01.13.21249759

**Authors:** Mengtian Du, Stacy L. Andersen, Thomas T. Perls, Paola Sebastiani

## Abstract

In recent years, there has been growing interest in the problem of model selection in the Bayesian framework. Current approaches include methods based on computing model probabilities such as Stochastic Search Variable Selection (SSVS) and Bayesian LASSO and methods based on model choice criteria, such as the Deviance Information Criterion (DIC). Methods in the first group compute the posterior probabilities of models or model parameters often using a Markov Chain Monte Carlo (MCMC) technique, and select a subset of the variables based on a prespecified threshold on the posterior probability. However, these methods rely heavily on the prior choices of parameters and the results can be highly sensitive when priors are changed. DIC is a Bayesian generalization of the Akaike’s Information Criterion (AIC) that penalizes for large number of parameters, it has the advantage that can be used for selection of mixed effect models but tends to prefer overparameterized models. We propose a novel variable selection algorithm that utilizes the parameters credible intervals to select the variables to be kept in the model. We show in a simulation study and a real-world example that this algorithm on average performs better than DIC and produces more parsimonious models.

## 1 Introduction

An important problem in statistics is choosing the model that best describes the data from a set of a priori plausible models. This problem is often reduced to variable selection from a set of explanatory variables assuming a general linear regression model, and Kadane and Lazar (2004) describe many criteria and search procedures that are applicable to modeling data from cross-sectional studies. In the context of analyzing longitudinal data, many variable selection methods have been proposed and extensively used for linear mixed effects models, including adaptation of the information criteria such as AIC (Akaike’s Information Criterion) and BIC (Bayesian Information Criterion), and shrinkage based methods such as LASSO (Least Absolute Shrinkage and Selection Operator). A review of some of these methods is provided by Muller et al. (2013).

In recent years, there also has been substantial interest in the variable selection problem within the Bayesian framework. There are two major approaches in the current Bayesian variable selection methods: methods based on computing model probabilities and methods based on computing model choice criteria. Many methods based on computing model probabilities are extensions of the “spike and slab” method first introduced by Mitchell and Beauchamp (1988). The general idea of this approach is to assign a prior distribution that mixes a point mass distribution at the null model and a diffuse uniform distribution elsewhere for each candidate variable. For each model, the posterior probabilities for both the vector of inclusion and coefficients are calculated through Markov Chain Monte Carlo (MCMC) techniques. The final subset of predictors are then selected based on a pre-specified threshold on the posterior probability. The Stochastic Search Variable Selection (SSVS) method proposed by George and McCulloch (1993) entails a specification of Bayesian mixture priors and uses the Gibbs sampling to identify the models with high posterior probability. A method suggested by Kuo and Mallick (1998) assigns an indicator posterior probability with value 1 if the posterior probability is sampled from the selected subset of predictors and value 0 when sampled from the full conditional distribution. The Gibbs Variable Selection (GVS) method suggested by Dellaportas et al. (2002) samples the parameter estimates from a mixture “pseudo-prior” that is concentrated around the posterior density of regression coefficients. Adaptive shrinkage methods such as Bayesian LASSO proposed by Park and Casella (2008) specify a prior directly over the regression coefficients to induce sparseness of the model. Rather than placing prior probabilities directly on the regression coefficients of the individual covariates, one could view the model as a whole and place prior directly on the number of covariates and their coefficients. Methods using this approach include reversible jump MCMC first proposed by Green (1995), and composite model (CMS) method introduced by Godsill (2001). Some of these approaches could achieve a relatively fast computation speed and good separation of variables as shown by O’Hara and Sillanpaa (2009). However, these methods rely on computing model posterior probabilities and are highly sensitive to the choice of priors so that the results may vary substantially with different prior distributions. The reviews by O’Hara and Sillanpaa (2009), Dellaportas et al. (2002), and George and McCulloch (1997) provide additional details and discussion.

There are limited options for Bayesian model selection criteria. A well-known criterion is the Deviance Information Criterion (DIC) that was proposed by Spiegelhalter et al. (2002). Similar to AIC, DIC penalizes for larger number of effective parameters in the model and models with smaller DIC are preferred. The criterion is implemented in OpenBUGS (BUGS, Bayesian inference Using Gibbs Sampling) and in JAGS (Just Another Gibbs Sampler). The latter implementation requires to run at least two parallel chains in the model. Recently Gelman et al. (2019), introduced a Bayesian version of R2, but this criterion needs to be evaluated.

Here we propose a variable selection algorithm that utilizes the parameters credible intervals to identify the variables to be retained in a model. The algorithm does not need “ad hoc” prior distributions of the regression parameters. It is computationally efficient and produces reasonable results. Vague conjugate priors can be assigned to the model coefficients without any need for tuning.

We will describe the algorithm in the next section. In Section 3 we describe the results of comprehensive simulations that show this algorithm on average produces more parsimonious models compared to DIC. In Section 4 we use the algorithm to test the hypothesis that genotypes of the *APOE* gene correlate with changes of cognitive function in a cohort of centenarians Sebastiani and Perls (2012). Conclusions and suggestions for future work are in Section 5.

## 2 Method

Consider a Bayesian model with outcome *y*_*i*_ for observation *i* (*i* = 1,…, *N*), a set of *h* + *p* possible predictors consisting of *h* variables *Z* = (*z*_1_, …, *z*_*h*_) to be kept in every model, and *p* candidate variables *X* = (*x*_1_, …, *x*_*p*_), where only an unknown subset of the *p* candidate variables may be relevant. The *Z* and *X* variables could either be main effects or interaction terms. We denote the set of possible parameter choices as *γ*_*m*_ = (*γ*_1_, …, *γ*_*p*_)^*T*^, where *γ*_*j*_ takes on values:

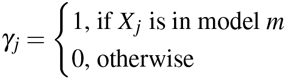

The regression model can then be expressed as

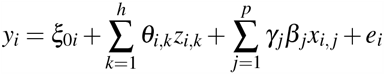

where *e*_*i*_ ∼ *N*(0, *σ*^2^) is the normally distributed error term with mean 0 and variance *σ*^2^ with Gamma prior. The term 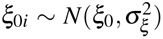 denotes a “random intercept” that we assume follows a normal distribution with mean *ξ*_0_ and variance, 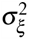 where *ξ*_0_ is the “fixed effect intercept” with mean 0 and variance 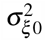. The parameter 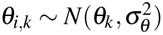 denotes the “random slope” for observation *i*, and *θ*_*k*_ denotes the “fixed effect” slope that we also assume follows a normal distribution with mean 0 and variance 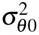 We assume the coefficients *β*_*j*_ are a priori independent and normally distributed with known mean and variance.

There are a total of |*γ*| =2^*p*^ plausible models based on the combinations of 0s and 1s in *γ*_*m*_, and we want to select the model that best describes the data. Denote by *C*_*α*_ = (*C*_*α*,1_, …,*C*_*α,p*_) the set of posterior credible intervals for the parameters *β* = (*β*_1_, …, *β*_*p*_) of the *p* candidate variables, where 1-*α* denotes the posterior coverage and each credible interval *C*_*α,j*_ consists of a lower bound *C*_*α,j,LB*_ and an upper bound *C*_*α,j,UB*_, *C*_*α,j*_ = (*C*_*α,j,LB*_,*C*_*α,j,UB*_). The CI algorithm proposed in this paper utilizes the credible intervals and its lower and upper bounds to perform variable selection.

The main idea behind the CI algorithm is to use the backward elimination method, first introduced by Marill and Green (1963) in the early 1960s, and a Bayesian metrics. In a traditional non-Bayesian multiple linear model, backward elimination begins with all candidate variables in the model and removes the least significant (largest p-value) variable one at a time, in an iterative way. In a Bayesian framework, we can utilize the posterior credible interval to help quantify the importance of each variable in the final model. We frame the variable selection problem as a hypothesis testing problem, in which rejecting the null hypothesis *H*_0_ : *β*_*j*_ = *β*_0,*null*_ leads to include the covariate *X*_*j*_ in the model. When the credible interval for the parameter *β*_*j*_ includes the null value, there are two alternative scenarios to consider. In the first scenario, the lower and upper bounds of the posterior credible interval are both far away from the null value, in other words, the null value falls well within the credible interval. In this case, we can say with “some confidence” that this variable is unlikely to be important. The second scenario is when either the lower or the upper bounds is close to the null value. In this case as more variables are dropped from the model, this variable will be more likely to be retained in the final model compared to the first scenario. In a non-Bayesian model, the second scenario could be interpreted as “borderline significant”. In each iteration step, we wish to remove the variable that is most likely to have a regression coefficient that satisfy the null hypothesis. Our observation, which is the rationale for the CI algorithm, is that for any credible interval that contains the null value, the minimum of the absolute value of the difference of the two bounds from the null value represents the importance of this variable and the CI algorithm identifies the variable with the minimum evidence against the null hypothesis to be removed at each iteration.

To perform variable selection, we first standardize all candidate variables to ensure that the regression coefficients are on the same scale. We start with the full model *Y* = *Zθ* + *Xβ*, which includes all *h* fixed variables *Z* and all *p* candidate variables *X*. In the first iteration, we obtain the posterior credible intervals *C*_*α,j*_(*j* = 1, …, *p*) for the parameters *β*_*j*_(*j* = 1, …, *p*) of each of the *p* candidate variables. If the credible interval of *x*_*j*_ does not include the null value (*β*_*j,null*_ ∉ *C*_*α,j*_), then *x*_*j*_ can remain in the model for the next iteration. Out of the *β*_*j*_’s with *β*_*j,null*_ ∈ *C*_*α,j*_, we remove from the model the variable *x*_*j*_ with max_*j ∈ p’*_ min(|*C*_*α,j,LB*_ − *β*_*j,null*_|, |*C*_*α,j,UB*_ − *β*_*j,null*_|), where *p*’ is the subset of parameters with *β*_*j,null*_ ∈ *C*_*α,j*_, and we repeat this process until all remaining candidate variables have *β*_*j,null*_ ∉ *C*_*α,j*_. A three-parameter example of this iteration process is illustrated in Figure 1.

**Fig. 1:**
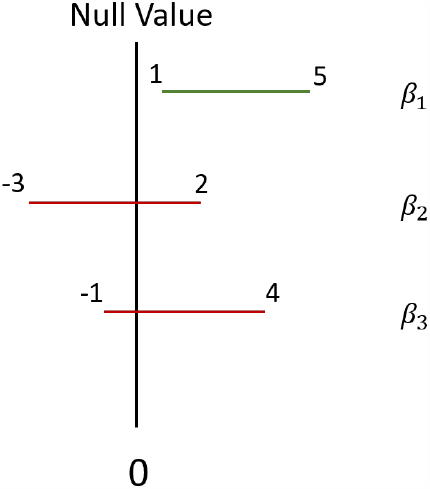
A three-parameter example of an iteration process of the CI algorithm.

In this example, there are three candidate variables for variable selection, namely *x*_1_, *x*_2_, and *x*_3_, with corresponding posterior estimates *β*_1_, *β*_2_, and *β*_3_ and 95% credible intervals *C*_0.95,1_ = (1, 5), *C*_0.95,2_ = (− 3, 2), and *C*_0.95,3_ = (− 1, 4). The null value is 0 for all *β*’s in this example. In the first iteration, the initial step is to identify the *β*’s with 95% CI that include 0 (0 ∈ *C*_0.95, *j*_), this case *β*_2_ and *β*_3_, while *β*_1_ has a 95% CI that does not include 0 and it can remain in the model for the next iteration. The second step is to identify which of *x*_2_ and *x*_3_ to remove by finding the maximum of minimum of the absolute value of the lower and upper bounds of the 95% CI (max_*j*∈*p’*_ {min(|*C*_*α,j,LB*_ − 0|, |*C*_*α,j,UB*_ − 0|)}). In this example, min(|*C*_0.95,2,*LB*_ − 0|, |*C*_0.95,2,*UB*_ − 0|) = min(| − 3|, |2|) = 2 for *β*_2_, and min(|*C*_0.95,3,*LB*_ − 0|, |*C*_0.95,3,*UB*_ − 0|) = min(| − 1|, |4|) = 1 for *β*_3_. Since 2 is greater than 1, we decide to remove *x*_2_ in this iteration. We will refit the model with only *x*_1_ and *x*_3_ in the next iteration and repeat the previous steps until all remaining candidate variables have posterior estimates 95% CI not including 0.

A method that has been extensively used and well implemented in Bayesian model selection is the Deviance Information Criterion (DIC) proposed by Spiegelhalter, et al. in 2002 Spiegelhalter et al. (2002). DIC is calculated as:

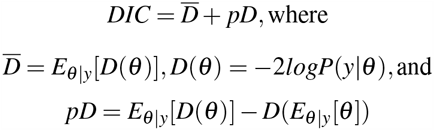

DIC is composed of two parts: the posterior mean of deviance 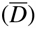 and the effective number of parameters (*pD*). Similar to the AIC, DIC penalizes for larger number of effective parameters in the model. We will be comparing the proposed credible interval algorithm and the DIC in the next section.

## 3 Empirical Evaluation

### 3.1 Simulation setup

We conducted a simulation study to evaluate the sensitivity and specificity of the proposed CI algorithm and to compare it with model selection based on DIC. Data used for simulation were generated based on the Digits Span Forward/Backward test from the Long Life Family Study (LLFS), which is a multi-center, longitudinal, family-based study of healthy aging and longevity Newman et al. (2011). The Digits Span test is a neuropsychological test that assesses auditory attention and working memory with score ranging from 0 to 14. The test was administered to approximately 4800 LLFS participants at enrollment, between 2006 and 2009. A second administration of the test occurred approximately 8 years after, in about 2500 participants, for a total N=7,289. We built two regression models of the test score, using predictors age at enrollment, follow-up time, gender, years of education, and familial longevity indicator (whether they were a member of a long-lived family or a spouse control). One model included only main effects, and the second model included the pairwise interactions between age at enrollment, follow-up time, and familial longevity indicator, sex, and education. We then used the two models to simulate datasets and evaluate the accuracy of the algorithm to identify the generating model. To perform variable selection using the CI algorithm, we used standardized covariates and started from the full model and went through the iterative steps to select the final model. The final selected model was checked against the simulated model to obtain level of concordance. The full model had the following form:

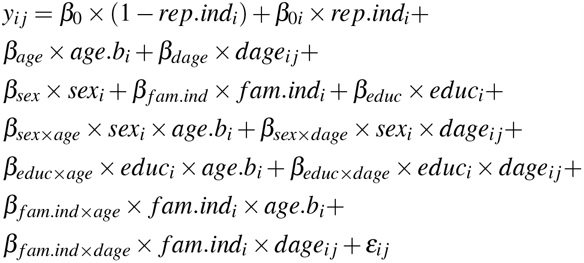

where *y*_*i j*_ represents the test score of the *i*^*t*^*h* individual at the *j*^*t*^*h* visit (*j*=0 for baseline and *j*=1 for follow-up) that we assumed normal distributed. To account for repeated measurements using a random intercept per study participant, we created an indicator variable *rep*.*ind* with value 1 if subject *i* had more than one measurements, and 0 otherwise. Covariates *age*.*b* and *dage* denoted age at baseline and follow-up time in years. The variable *sex* was a binary variable with value 1 for males and 0 for females, and variable *educ* was an ordinal variable with values 0-17 that approximated years of education. The variable *f am*.*ind* was an indicator variable with value 1 if subject *i* was a member of a long-lived family and 0 if subject i was a spouse control. The random intercept term *β*_0*i*_ was assigned a normal prior distribution with mean *β*_0_ and precision parameter *τ*, which had Gamma distribution with both shape and scale parameters equal to 1. All other covariates were assigned normal prior distributions with mean 0 and precision 0.1.

A “mismatch” occurred when the CI algorithm falsely detected an interaction term in the analysis of the data generated from the main effects model (a false positive) or failed to detect a real interaction in the analysis of the data generated from the full model (a false negative). We run the simulations with four different sample sizes: 500, 1000, 5000, and the largest sample size we had, 7289, with 100 replications for each sample size. As a comparison to the CI algorithm, we also performed model selections using DIC. In each simulation setting (main effect model and full model) and with each sample size, we ran two parallel chains of the MCMC in the R package JAGS, and computed the DIC for each of the 2^*p*^=64 models and selected the final model with the smallest DIC. All Bayesian models in the CI algorithm were ran with 2,000 adaptions and 5,000 iterations, and all Bayesian models used in the DIC method were ran with 2,000 adaptions and 1,000 iterations to reduce the computing time.

### 3.2 Simulation results

The results of the simulation are shown in Table 1 and Figure 2. When the data were generated from the full model, with sample sizes 1,000, 5,000, and 7,289, both the CI algorithm and DIC detected the correct model with 100% accuracy. With the smallest sample size 500, the CI algorithm detected the correct model with 74% accuracy and 26% 1-mismatch rate (only five out of the six interaction terms were detected), and DIC detected the correct model with 87% accuracy and 13% 1 mismatch rate. This is the only scenario where the DIC performed better than the CI algorithm in the full model setting. When the data were generated from the model with main effects only, the CI algorithm detected the correct model with 100% accuracy with sample sizes 500, 1,000, and 7,289, and with 99% accuracy and 1% 2-mismatch rate (falsely detect two interaction terms) with sample size 5,000. However, DIC only detected the correct model with 37%, 25%, 9% and 3% accuracy in the sample sizes 500, 1,000, 5,000, and 7,289, respectively. As shown in Table 1 and Figure 2, DIC tended to falsely select interaction terms which are more concentrated with one, two, and three mismatches. The simulation results suggest that the CI algorithm favors more parsimonious models than DIC. This property may lead to miss important effects with small sample sizes. DIC appears to favor models with more redundant parameters and may be a better choice for predictive modeling. However, performing a full search using DIC is only computationally feasible with a relatively small number of parameters. In a model with p candidate variables, calculating the DIC for 2^*p*^ models could be extremely computationally intensive as p gets large. Although one can also perform variable selection using a backward approach with DIC, it is still very computationally demanding because it requires running multiple chains for each model.

**Table 1:**
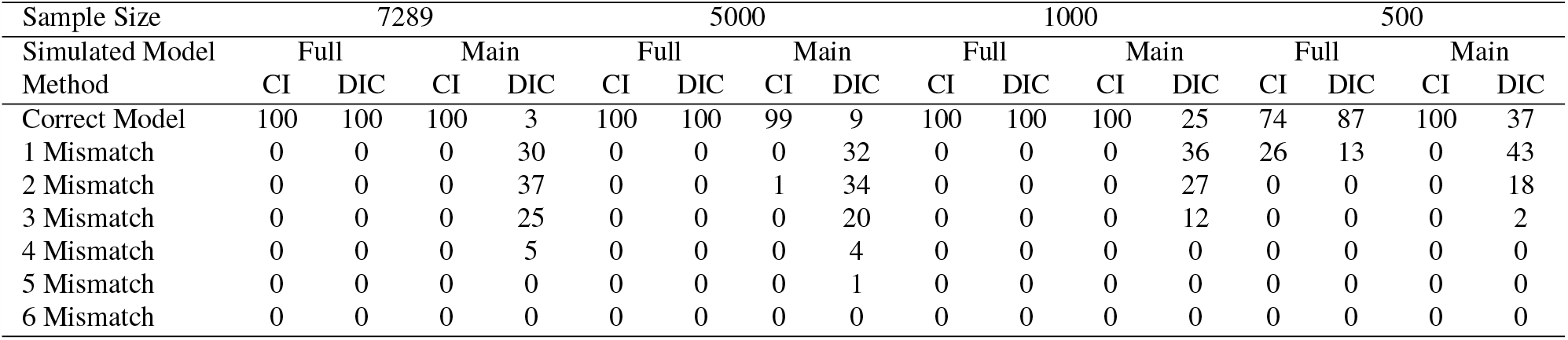
Simulation results comparing the CI algorithm and DIC.

**Fig. 2:**
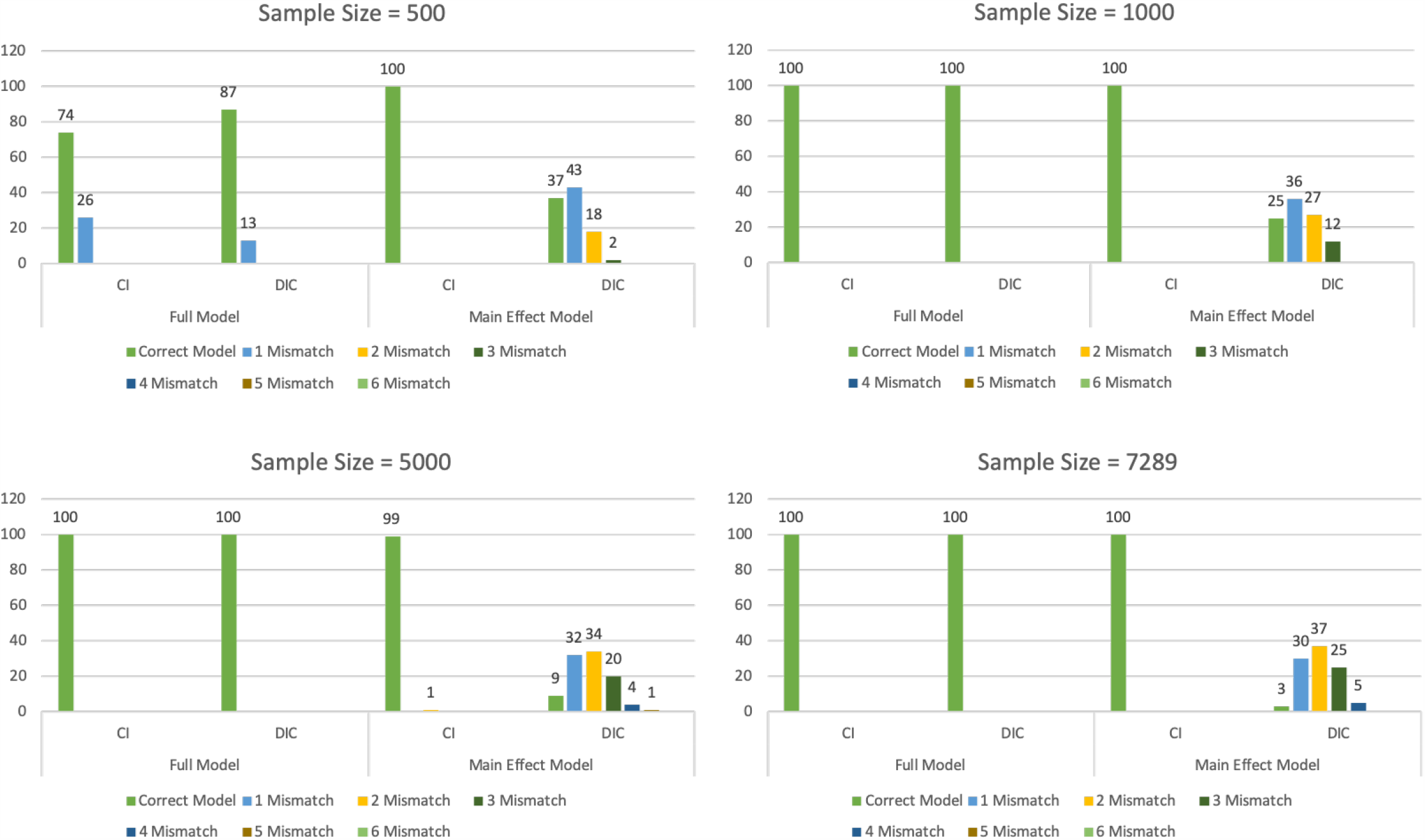
Simulation results for both full model and main effect model.

All analyses were run in R3.5.1 using the rjags package version 4-6.

## 4 Application: Testing the Effect of *APOE* Alleles in Cognitive Decline of Centenarians

### 4.1 Background

There is growing evidence that the rate of change of cognitive function is genetically regulated by the gene *APOE*: a well-established risk factor for Alzheimer’s disease. One variant of this gene is associated with extreme human longevity and possibly with slower decline of cognitive function, as shown by Sebastiani et al. (2019); Kim et al. (2017). We tested this hypothesis using data from the New England Centenarian Study (NECS): a longitudinal study of centenarians born between 1880 and 1910 and their family members that is described by Sebastiani and Perls (2012). Study participants have been enrolled since 1995 and are followed-up annually, until death, to assess their health status and their physical and cognitive functions. The latter is assessed by administering the Information-Memory-Concentration (IMC) portion of the Blessed Dementia Scale (see work by Blessed et al. (1968)). The total memory score of the IMC ranges from 0 to 37, where 33 or higher indicating normal cognition; 27 to 32 indicating mild cognitive impairment; 21 to 26 indicating moderate impairment; and 21 or lower indicating severe cognitive impairment (Terry et al. (2008)). Genotype data of the *APOE* gene have been generated as described in Sebastiani et al. (2019) using a combination of genome-wide genotype data that were generated with Illumina arrays, and imputation. We generated *APOE* alleles e2, e3 and e3 from combinations of the genotypes of the single nucleotide polymorphisms (SNPs) rs7412 and rs429358 as described in Sebastiani et al. (2019). We then analyzed the total memory score as a function of the *APOE* variants to test the hypothesis that different genotypes of this gene correlate with different rate of cognitive decline at extreme old age.

### 4.2 Analysis

We stratified the 485 NECS participants with *APOE* genotype information and at least one IMC score (excluding participants with e2e4 genotype) into three groups: the “APOE2 group” (N=117), which comprised carriers of the genotypes e2e2 or e2e3; the “APOE3 group” (N=330), which comprised carriers of e3e3; and the “APOE4 group” (N=38), which comprised carriers of e3e4 (genotype e4e4 was not available in this study sample). The “APOE3 group” was used in the analysis as the reference group because it is the most common genotypes in whites. A summary of demographic information is shown in Table 2. Age at enrollment ranged from 91 to 113 years with mean of 103.3 years (SD: 4.5). There were no significant differences in age at enrollment, average follow-up time, gender, education and score at first assessment comparing the APOE2 group and the APOE4 group to the APOE3 group.

**Table 2:** Demographic characteristics and test score of 485 NECS study participants.

|  | APOE2(e2e2, e2e3) | APOE3(e3e3) | APOE4(e3e4) | p-value (t-test,<br>comparing<br>APOE2 and<br>APOE3) | p-value (t-test,<br>comparing<br>APOE4 and<br>APOE3) |
| --- | --- | --- | --- | --- | --- |
| N(%) | 117(24.1%) | 330(68%) | 38(7.8%) |  |  |
| Age at Enrollment, mean(SD), years | 103.5(4.6) | 103.3(4.4) | 102.4(5) | 0.66 | 0.29 |
| Follow-up, mean(SD), years | 1.8(2) | 1.7(2.1) | 1.9(2.3) | 0.89 | 0.67 |
| Gender, male(%) | 22(18.8%) | 85(25.8%) | 9(23.7%) | 0.11 | 0.78 |
| Education, years | 19.2(13.3) | 18.1(12.8) | 17.6(13.3) | 0.48 | 0.81 |
| Total Memory Score at first assessment (SD) | 25.1(8.3) | 25(8.6) | 24.4(10.2) | 0.9 | 0.72 |

To test the association between the *APOE* genotype and the rate of memory decline, we compared the effects of APOE2 and APOE4 to APOE3 separately. In the APOE2 analysis, we subset our sample to participants in the APOE2 group and the APOE3 group, and a new APOE variable was created to take on value 1 if participant carried at least one cope of the e2 allele, and value 0 if participant had genotype e3e3. Similarly, the APOE4 analysis included only participants from the APOE4 group and the APOE3 group, and a new APOE variable was created to take on value 1 if participant carried at least one e4 allele, and value 0 if participant had genotype e3e3. We used a Bayesian hierarchical model with random intercept to model the association between the outcome composite memory score and the *APOE* groups, adjusting for other factors. The full model for both APOE2 and APOE4 analyses had the following form:

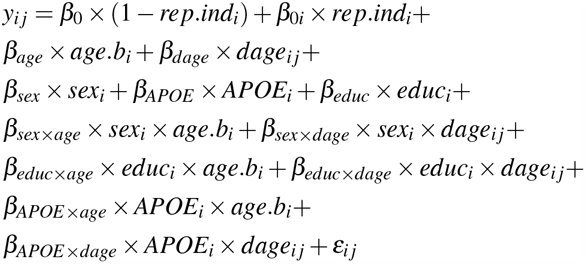

where *y*_*i j*_ represented the Blessed composite memory score for the *i*^*t*^*h* individual and the *j*^*t*^*h* visit (*j*=1 for baseline visit and *j*=2 for follow-up visit). To account for repeated measurements, the intercept of this model *β*_0_ × (1 − *rep*.*ind*_*i*_) + *β*_0*i*_ × *rep*.*ind*_*i*_ had both a fixed effect *β*_0_ with normal prior distribution of mean 0 and precision 0.1, and a random effect *β*_0*i*_ with normal prior distribution of mean *β*_0_ and precision parameter *τ*, which had prior distribution Gamma with both shape and scale parameters equal to 1. The indicator *rep*.*ind*_*i*_ was defined as described earlier to indicate whether individual *i* had more than one measurement (*rep*.*ind*_*i*_=1 if the *i*^*t*^*h* individual had more than one measurement, and 0 otherwise). The error term *ε*_*i j*_ was assumed to have Normal distribution with 0 mean and variance that was a priori followed an inverse Gamma distribution (both shape and scale parameters equal to 1). The covariates *age*.*b* and *dage* represented age at baseline and follow-up time in years. The sex variable was coded to have value 1 for male and 0 for female, and educ was an ordinal variable taking on values 0-17 that approximates years of education. The APOE variable was created, as described above, to take on value 1 to indicate individual i had at least one copy of e2 or e4 allele, and 0 if their *APOE* genotype was e3e3. There were six interaction terms in the full model, including interaction terms between sex, education, *APOE*, and age at baseline and follow-up time in years. The two interactions *β*_*APOE*×*age*_ and *β*_*APOE*×*dage*_ represented the effect of *APOE* on the cross-sectional and longitudinal effect of age.

To perform the model selection and obtain final model estimates, we first standardized all variables in the model. During the model selection process, all main effect terms were always kept in the model, and interaction terms were considered as the candidate variables for selection. We started with the full model in the first iteration and removed the interaction term with minimum evidence against the null hypothesis (*β* = 0) identified by the CI algorithm. This process was repeated until all interaction terms remaining in the model had credible intervals not including the null value, or until all interaction terms were removed from the model. To provide the final model parameter estimates, the posterior estimates of main effects were scaled back by divide by the standard deviation of corresponding variable, and the posterior estimate of interaction terms were scaled back by divide by the product of standard deviations of the corresponding variables.

All analyses were conducted in R 3.5.1 using the rjags package version 4-6.

### 4.3 Results

Parameter estimates of the final models are shown in Table 3. In neither analysis, the CI algorithm detected interactions between the *APOE* alleles and either age at enrollment or followup time. In the APOE2 analysis (comparing the APOE2 group to the APOE3 group), both age at enrollment and follow-up time were negatively associated with Blessed total memory score (age effect = −0.8, 95%CI: −0.94, 0.66; follow-up time effect = -1.11, 95%CI: -1.36, −0.87). This suggests that for every one year increase in age at enrollment and follow-up time, the Blessed total memory score was expected to decrease by 0.8 points and 1.11 points, respectively. Years of education and having at least one copy of the e2 allele did not appear to have an effect on the total memory score. Particularly, carriers of 1 or more copies of the e2 allele had an estimated increase of the Blessed memory score of 0.37, although the posterior probability that this effect is *>* 0 was only 0.12. Similarly, in the APOE4 analysis (comparing the APOE4 group to the APOE3 group), age at enrollment and follow-up time were negatively associated with the total memory score (age effect = −0.82, 95%CI: - 0.97, −0.67; follow-up time effect = -1.15, 95%CI: -1.42, - 0.88). Years of education and having at least one copy of the e4 allele did not appear to have a strong effect on the total memory score: carriers of 1 or more copies of the e4 allele had an estimated decrease of the Blessed memory score of 1.78, although the posterior probability that this effect is *<* 0 was only 0.05. These results suggest that there is no strong association between *APOE* genotype and the Blessed total memory score in very old people, and that there is no strong difference in rate of change of Blessed total memory score among *APOE* genotype groups. We have also performed variable selection using DIC as a comparison to the CI algorithm. In the APOE2 and APOE4 analyses, DIC has selected models with four interaction terms and two interaction terms respectively, and all interaction terms had parameter estimates 95% credible intervals including 0.

**Table 3:** Parameter estimates of the New England Centenarian Study data.

|  | APOE2 | APOE4 |
| --- | --- | --- |
| age | -0.8(-0.94,-0.66) | -0.82(-0.97,-0.67) |
| dage | -1.11(-1.36,-0.87) | -1.15(-1.42,-0.88) |
| sex, male | 2.48(1.02,3.89) | 2.21(0.65,3.76) |
| educ | -0.01(-0.06,0.03) | -0.04(-0.09,0.01) |
| APOE | 0.37(-0.94,1.66) | -1.78(-3.92,0.37) |
| educ*age | - | - |
| educ*dage | - | - |
| sex*age | - | - |
| sex*dage | - | - |
| APOE*age | - | - |
| APOE*dage | - | - |

## 5 Conclusion

In this article, we proposed the CI algorithm: a novel approach to perform Bayesian variable selection utilizing the posterior credible intervals. Inspired by the backward elimination variable selection method in linear regression models, this algorithm removes candidate variables one at a time by quantifying and comparing the strength of the association of possible predictors with the outcome. We conducted a comprehensive simulation to assess the sensitivity and specificity of the algorithm and the simulation suggests that the algorithm is accurate with relatively large samples, and compared to DIC tends to produce more parsimonious models with smaller false positive rate.

There are a few advantages of our proposed CI algorithm comparing to some variable selection methods based on computing model probabilities and methods based on computing model choice criteria such as DIC. First, the CI algorithm does not require to specific prior distributions on the model coefficients and the inclusion probabilities of each predictor, or a posterior threshold to decide whether or not to include a predictor. Secondly, our method yields interpretable results. The final selected model all have 95% credible intervals not containing the null value. When implemented with real world data, these results could help derive practical meaningful interpretations. Thirdly, although in this paper we illustrated our proposed algorithm using a mixed effects model with random intercepts, it can be extended and applied to a more generalized form of linear models. For variables with random effects, we can still perform variable selection on the fixed part of the random effects. And lastly, the CI algorithm is computationally efficient and the largest number iterations (number of Bayesian models to run) is equal to the number of candidate variables. Out of the methods discussed above, DIC is the most computationally intensive since it requires to run at least two parallel MCMC chains of all possible 2^*p*^ models. “Spike and slab” based methods require to run long MCMC chains to search over model space, which also could be computationally demanding. However, computation load of our algorithm could still get immense in the case of high-dimensional data where *p* ≫ *n*. Bondell and Reich (2012) proposed a Bayesian variable selection approach utilizing joint credible regions that can be suited for the high-dimensional case, though this approach yields higher false positive rate in the low-dimensional setting compared to our proposed algorithm based on their simulation study.

Overall, our proposed method performs well with reasonable number of parameters. It yields results with parsimonious parameters with practical meaning and can achieve computational efficiency. In addition to the application example in this paper, we have also implemented this proposed algorithm in analyzing the association between familial longevity and cognitive decline, and the association between *APOE* allele and cognitive decline in the Long Life Family Study (LLFS) Andersen et al. (2020); Du et al. (2020).

## Data Availability

Data is available via dbGaP (dbGaP Study Accession: phs000397.v1.p1)

## Conflict of interest

The authors declare that they have no conflict of interest.

---

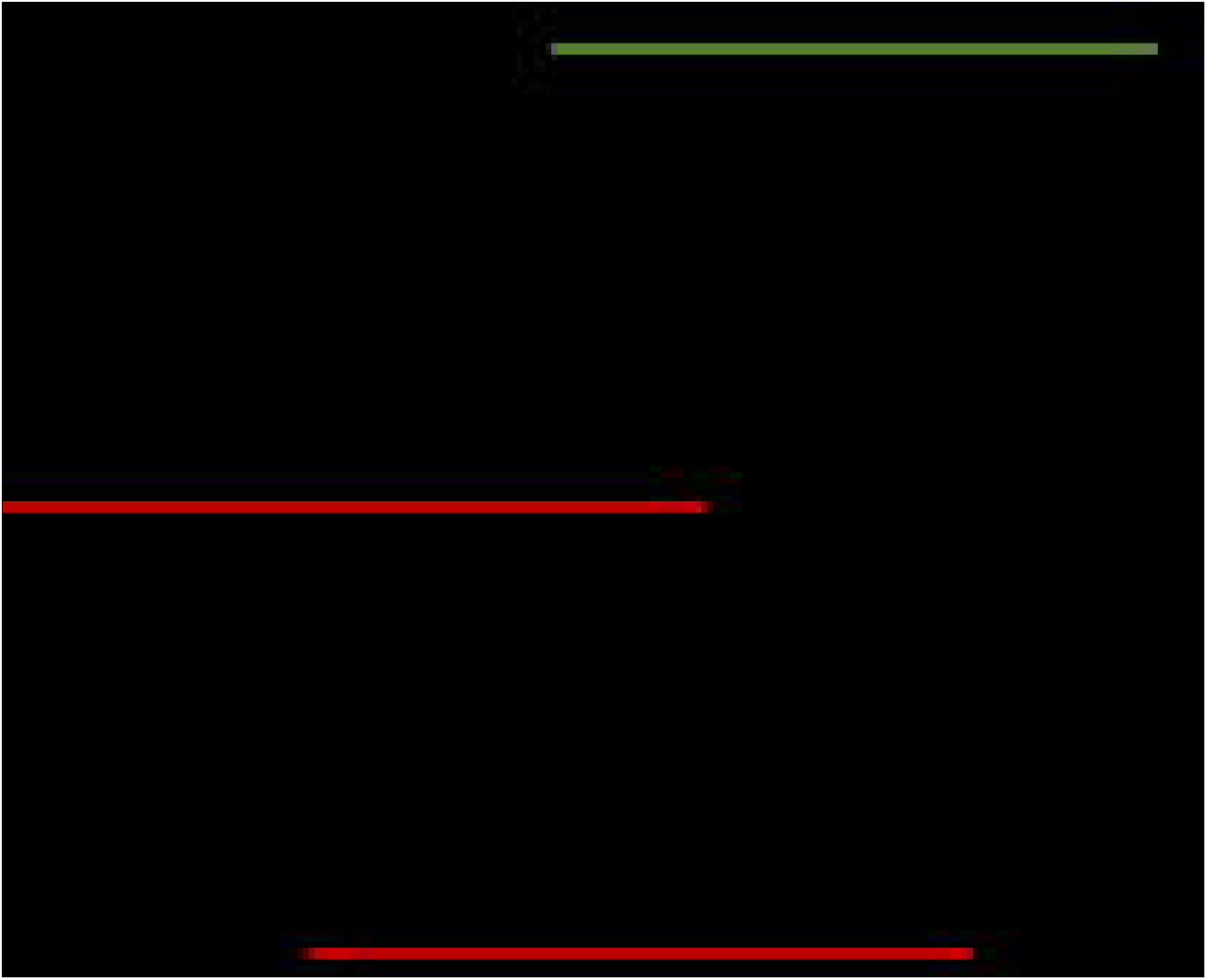

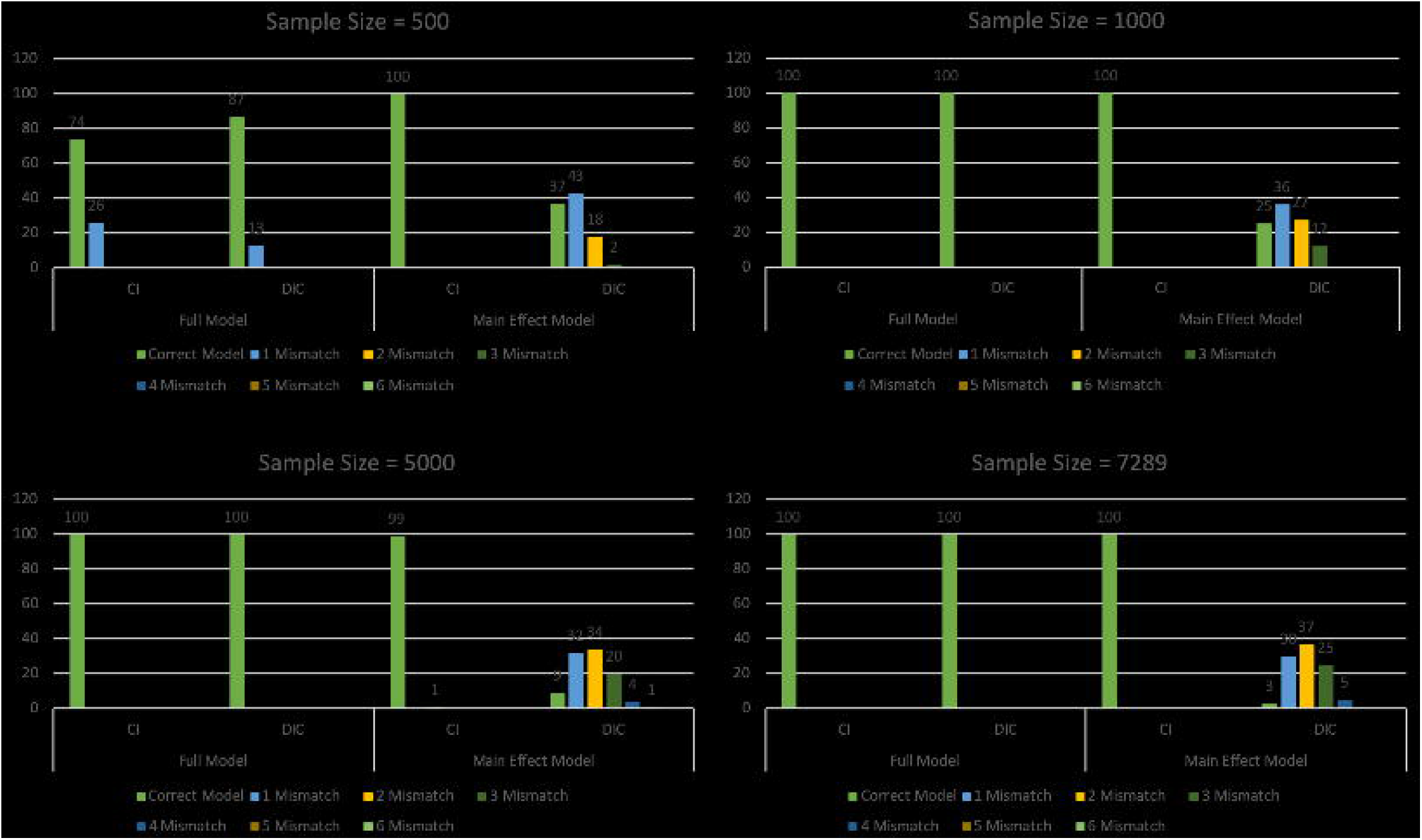

## Notes

### Competing Interest Statement

The authors have declared no competing interest.

### Funding Statement

This work was supported by the NIH: R01AG061844 to PS, K01 AG057798 to SA; TP; U19AG063893 to PS, TP.

### Author Declarations

The IRB from Washington University St Luis approved the protocol

## References

Andersen SL, D. M, Cosentino S, Schupf N, Rosso AL, Perls TT, Sebastiani P (2020) Slower decline in processing speed is associated with familial longevity

Blessed G, Tomlinson BE, Roth M (1968) The association between quantitative measures of dementia and of senile change in the cerebral grey matter of elderly subjects. Br J Psychiatry 114(512):797–811, DOI 10.1192/bjp.114.512.

Bondell HD, Reich BJ (2012) Consistent high-dimensional bayesian variable selection via penalized credible regions. Journal of the American Statistical Association 107(23482517):1610–1624

Dellaportas P, Forster JJ, Ntzoufras I (2002) On bayesian model and variable selection using mcmc. Statistics and Computing 12(1):27–36, DOI 10.1023/A:1013164120801

Du M, Andersen SL, Schupf N, Feitosa MF, Barker MS, Perls TT, Sebastiani P (2020) Association between apoe alleles and change of neuropsychological tests in the long life family study

Gelman A, Goodrich B, Gabry J, Vehtari A (2019) R-squared for bayesian regression models. The American Statistician 73(3):307–309, DOI 10.1080/00031305.2018.1549100

George EI, McCulloch RE (1993) Variable selection via gibbs sampling. Journal of the American Statistical Association 88(423):881–889, DOI 10.1080/01621459.1993.10476353

George EI, McCulloch RE (1997) Approaches for bayesian variable selection. Statistica Sinica 7(2):339–373

Godsill SJ (2001) On the relationship between markov chain monte carlo methods for model uncertainty. Journal of Computational and Graphical Statistics 10(2):230–248

Green PJ (1995) Reversible jump markov chain monte carlo computation and bayesian model determination. Biometrika 82(4):711–732, DOI 10.2307/2337340

Kadane JB, Lazar NA (2004) Methods and criteria for model selection. Journal of the American Statistical Association 99(465):279–290, DOI 10.1198/016214504000000269

Kim YJ, Seo SW, Park SB, Yang JJ, Lee JS, Lee J, Jang YK, Kim ST, Lee KH, Lee JM, Lee JH, Kim JS, Na DL, Kim HJ (2017) Protective effects of apoe e2 against disease progression in subcortical vascular mild cognitive impairment patients: A three-year longitudinal study. Sci Rep 7(1):1910, DOI 10.1038/s41598-017-02046-y 5

Kuo L, Mallick B (1998) Variable selection for regression models. Sankhyā: The Indian Journal of Statistics, Series B (1960-2002) 60(1):65–81 2

Marill T, Green D (1963) On the effectiveness of receptors in recognition systems. IEEE Transactions on Information Theory 9:11–17, DOI 10.1109/TIT.1963.1057810 3

Mitchell TJ, Beauchamp JJ (1988) Bayesian variable selection in linear regression. Journal of the American Statistical Association 83(404):1023–1032, DOI 10.1080/01621459.1988.10478694 2

Muller S, Scealy JL, Welsh AH (2013) Model selection in linear mixed models. Statist Sci 28(2):135–167, DOI 10.1214/12-STS410 2|

Newman AB, Glynn NW, Taylor CA, Sebastiani P, Perls TT, Mayeux R, Christensen K, Zmuda JM, Barral S, Lee JH, Simonsick EM, Walston JD, Yashin AI, Hadley E (2011) Health and function of participants in the long life family study: A comparison with other cohorts. Aging (Albany NY) 3(1):63–76, DOI 10.18632/aging.100242 4|

O’Hara RB, Sillanpaa MJ (2009) A review of bayesian variable selection methods: what, how and which. Bayesian Anal 4(1):85–117, DOI 10.1214/09-BA403 2|

Park T, Casella G (2008) The bayesian lasso. Journal of the American Statistical Association 103(482):681–686, DOI 10.1198/016214508000000337 2|

Sebastiani P, Perls TT (2012) The genetics of extreme longevity: lessons from the new england centenarian study. Front Genet 3:277, DOI 10.3389/fgene.2012.002773, 5|

Sebastiani P, Gurinovich A, Nygaard M, Sasaki T, Sweigart B, Bae H, Andersen SL, Villa F, Atzmon G, Christensen K, Arai Y, Barzilai N, Puca A, Christiansen L, Hirose N, Perls TT (2019) Apoe alleles and extreme human longevity. J Gerontol A Biol Sci Med Sci 74(1):44–51, DOI 10.1093/gerona/gly174 5|

Spiegelhalter DJ, Best NG, Carlin BP, Van Der Linde A (2002) Bayesian measures of model complexity and fit. Journal of the Royal Statistical Society: Series B (Statistical Methodology) 64(4):583–639, DOI 10.1111/1467-9868.003532, 4|

Terry DF, Sebastiani P, Andersen SL, Perls TT (2008) Disentangling the roles of disability and morbidity in survival to exceptional old age. Arch Intern Med 168(3):277–83, DOI 10.1001/archinternmed.2007.75 5|

